# Comparison of the classification of HER2 from whole-slide images between pathologists and a deep learning model

**DOI:** 10.1101/2023.03.29.23287897

**Authors:** Masayuki Tsuneki, Makoto Abe, Fahdi Kanavati

**Affiliations:** Medmain Research, Medmain Inc., Fukuoka, 810-0042, Japan; Department of Pathology, Tochigi Cancer Center, 4-9-13 Yohnan, Utsunomiya 320-0834, Tochigi, Japan

## Abstract

HER2 (human epidermal growth factor receptor 2) is a protein that is found on the surface of some cells, including breast cells. HER2 plays a role in cell growth, division, and repair, and when it is overexpressed, it can contribute to the development of certain types of cancer, particularly breast cancer. HER2 overexpression occurs in approximately 20% of cases, and it is associated with more aggressive tumor phenotypes and poorer prognosis. This makes its status an important factor in determining treatment options for breast cancer. While HER2 expression is typically diagnosed through a combination of immunohistochemistry (IHC) and/or fluorescence in situ hybridization (FISH) testing on breast cancer tissue samples, we sought to determine to what extent it is possible to diagnose from H&E-stained specimens. To this effect we trained a deep learning model to classify HER2-positive image patches using a dataset of 10 whole-slide images (5 HER2-positive, 5 HER2-negative). We evaluated the model on a different test set consisting of patches extracted from 10 WSIs (5 HER2-positive, 5 HER2-negative), and we compared the performance against two pathologists on 100 512×512 patches (50 HER2-positive, 50 HER2-negative). Overall, the model achieved an accuracy of 73% while the pathologists achieved 58% and 47%, respectively.

## 1 Introduction

The HER-2/neu (c-erbB-2) is an oncogene that encodes a transmembrane glycoprotein with tyrosine kinase activity known as p185, which belongs to the family of epidermal growth factor receptors [1, 2]. Overexpression of HER-2 in breast cancer has been associated with poor overall survival and has been shown preclinically to enhance malignancy and the metastatic phenotypes [1]. HER2 overexpression can be identified by immunohistochemistry (IHC) or fluorescence in situ hybridization (FISH) on histopathological specimens [3]. It has been revealed that repression of HER-2 suppresses the malignant phenotypes of HER-2-overexpressing cancer cells, which strongly suggests that HER-2 may serve as an excellent target for anticancer agents specific for HER-2-overexpressing cancer cells [1]. According to the American Society of Clinical Oncology (ASCO) and College of American Pathologists (CAP) guidelines in 2018 [4], surgical pathologists select invasive breast cancer patients to be treated with anti-HER2 drugs. IHC 0 (negative) is defined as invasive breast cancer with faint membrane staining observed in less than 10% of cancer cells. IHC 1+ (negative) is defined as invasive breast cancer with faint membrane staining observed in over 10% of cancer cells. IHC 2+ (equivocal) is defined as invasive breast cancer with weak to moderate complete membrane staining observed in over 10% cancer cells. IHC 3+ (positive) is defined as invasive breast cancer with strong complete membrane staining observed in over 10% cancer cells [4]. According to the guideline, in this study, we defined IHC 0 and IHC 1+ as negative and IHC 3+ as positive.

In this short study we aimed to investigate the feasibility of diagnosing HER2 expression from H&E-stained specimens, as the current standard practice involves using IHC and/or FISH testing on breast cancer tissue samples. We developed a deep learning model to classify HER2-positive image patches using a dataset of 10 whole-slide images, including 5 HER2-positive and 5 HER2-negative cases. We then evaluated the performance of the model on a separate test set consisting of patches extracted from 10 whole-slide images (5 HER2-positive, 5 HER2-negative), and compared it to the performance of two pathologists who assessed 100 512×512 patches (50 HER2-positive, 50 HER2-negative). The results of our study showed that the deep learning model achieved an accuracy of 73% in HER2 classification, which outperformed the pathologists’ accuracy of 58% and 47%, respectively. On the entire set of test patches the model achieve an ROC AUC of 0.744 [0.712-0.767]. These findings suggest that it may be possible to diagnose HER2 expression from H&E-stained specimens using deep learning models, which could potentially improve the efficiency and accuracy of HER2 testing in clinical practice.

## 2 Method

### 2.1 Dataset

In this retrospective study, 10 HER2 positive breast cancer, 10 HER2 negative breast cancer were collected from the surgical pathology files of Kamachi Group Hospitals (Wajiro and Shinkomonji hospitals, Fukuoka, Japan) after histopathological review of all specimens by surgical pathologists. We collected both H&E stained and HER2 immunohistochemical stained glass slides from hospitals. The glass slides were digitised into WSIs at magnification x20 using a Leica Aperio AT2 Digital Whole Slide Scanner (Leica Biosystems, Tokyo, Japan) was used to scan the slides at a magnification of x20. The diagnosis of each specimen was extracted from the medical reports. In the training set, we used 5 HER2 positive WSIs (5 IHC 3+), 5 HER2 negative WSIs (four IHC 0 and one IHC 1+). In the test set, we used 5 HER2 positive WSIs (5 IHC 3+) and 5 HER2 negativWSIs (four IHC 0 and one IHC 1+).

One pathologist manually annotated HER2 positive and HER2 negative breast cancer areas by reviewing HER2 immunohistochemical WSIs (Figure 2.1). We set two annotation labels: HER2 positive and HER2 negative. The average annotation time per WSI was about 15 min.

From the annotations areas in the training set, we extracted 95,561 patches for HER2 positive and 136,132 for HER2 negative. In the test set, we extracted 147,837 HER2 positive and 124,518 HER2 negative. The patch size was 512×512 at magnification x20 with an overlap stride of 128×128.

**Figure 1:**
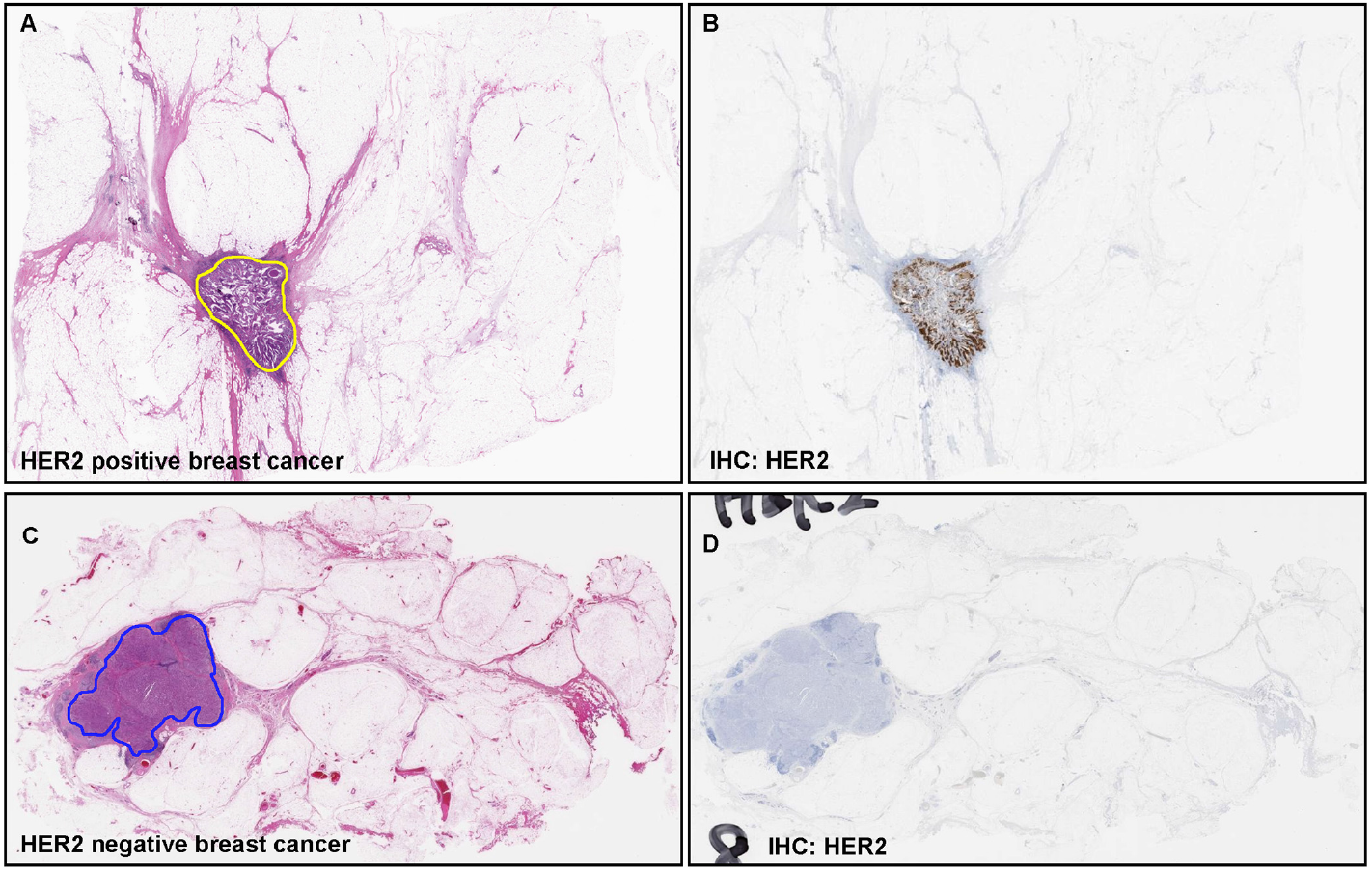
Annotation overview.

### 2.2 Deep learning model

The deep learning model was trained using a weakly-supervised training method exactly as described in previous works[5, 6]. To refer the reader to the referenced publications for training details. We performed tissue detection using Otsu’s thresholding method [7] by excluding the white background. We then extracted tiles only from the tissue regions. During prediction, we extracted tiles from the entire tissue regions using a sliding window with a fixed-size stride.

## 3 Results

### 3.1 Patch-level evaluation

We first evaluated the performance of the model on a test set of 512×512 patches consisting of 147,837 HER2 positive and 124,518 HER2 negative. The model achieved an ROC AUC of 0.744 [0.712-0.767]. We summarise the results in Fig. 3.1.

**Figure 2:**
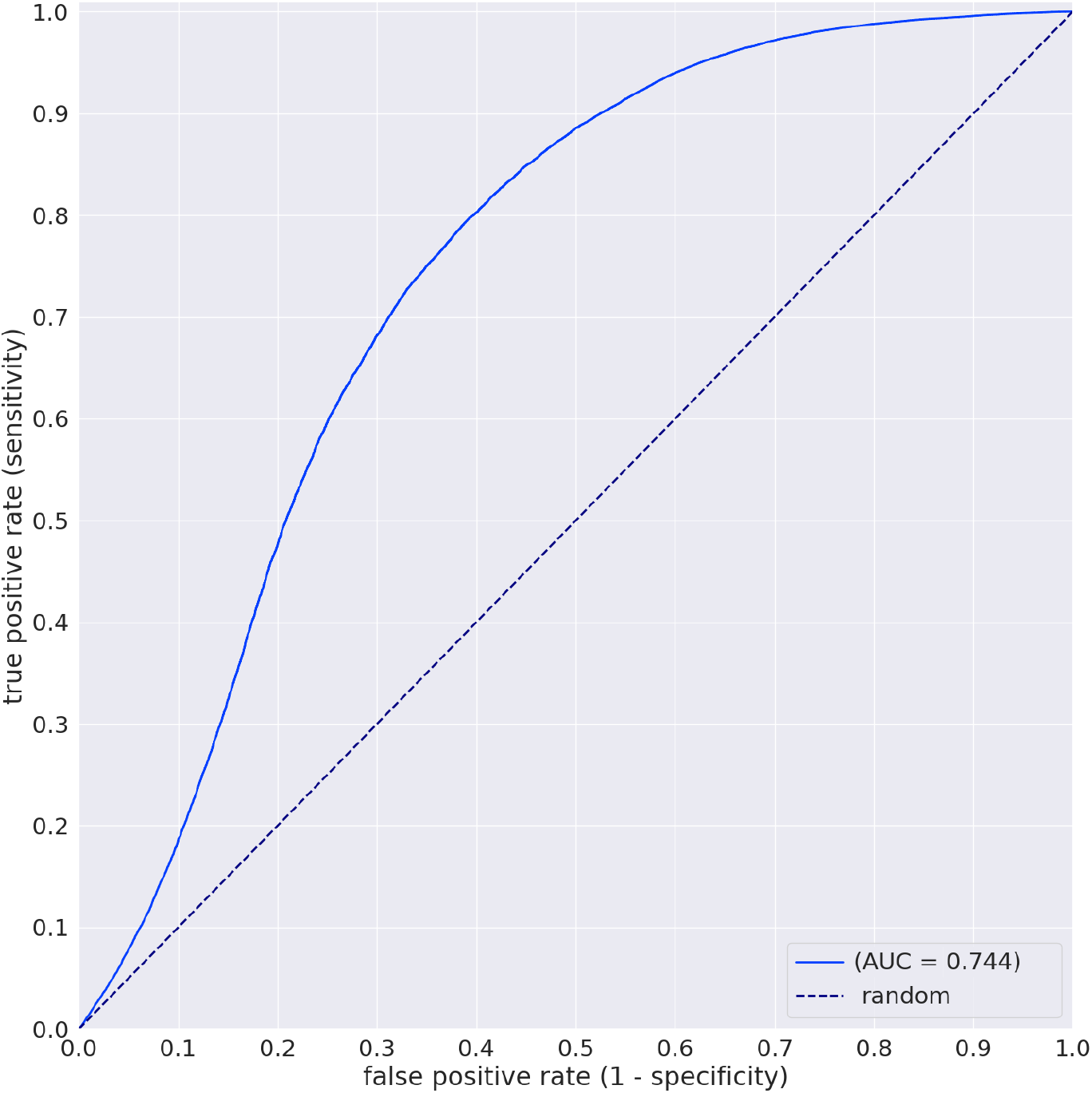
ROC for classifying HER2 positive as evaluated on the patches in the test set.

### 3.2 Comparaison against two pathologists

In addition to evaluating the performance of our deep learning model, we also assessed the ability of two pathologists to detect HER2 protein expression from WSI patches. The pathologists independently reviewed 100 512×512 patches (50 HER2 positive, 50 HER2 negative) that were randomly extracted from 10 WSIs, and they provided a binary classification (HER2 positive or HER2 negative) for each patch. This evaluation was carried out two months after the annotation process to allow enough time for the pathologists to not easily recall particular cases. The pathologists achieved 58% and 47%, respectively, while the model achieved an accuracy of 73%.

## 4 Conclusion

The results of the study showed that the deep learning model performed better than two pathologists in classifying HER2 protein expression from patches extracted from breast cancer WSIs.

Even though H&E slides are not the primary way used for diagnosing HER2 protein expression, the study demonstrated that a deep learning model could potentially achieve better accuracy at it compared to pathologists, meaning that the model is most likely picking up on subtle patterns which are harder to pick up by pathologists that are not typically used to detecting HER2 protein expression from histopathology images. The results suggest that deel learning models can be a useful tool for pathologists, particularly in settings where access to expert pathologists is limited, and they can help improve the consistency of detection.

## Data Availability

Due to specific institutional requirements governing privacy protection, datasets used in this study are not publicly available.

## Acknowledgements

We are grateful for the support provided by Dr. Shigeo Nakano at Kamachi Group Hospitals (Fukuoka, Japan). We thank pathologists who have been engaged in reviewing cases, performing annotation, and clinicopathological discussion for this study. This study is based on results obtained from a project, JPNP14012, subsidized by the New Energy and Industrial Technology Development Organization (NEDO).

## Ethics approval and consent to participate

The experimental protocol was approved by the ethical board of Kamachi Group Hospitals (Wajiro and Shinkomonji hospitals, Fukuoka, Japan) (No. 173). All research activities complied with all relevant ethical regulations and were performed in accordance with relevant guidelines and regulations in the all hospitals mentioned above (Wajiro and Shinkomonji hospitals, Fukuoka, Japan). Informed consent to use histopathological samples and pathological diagnostic reports for research purposes had previously been obtained from all patients prior to the surgical procedures at all hospitals, and the opportunity for refusal to participate in research had been guaranteed by an opt-out manner.

## Funding

This study is based on results obtained from a project, JPNP14012, subsidized by the New Energy and Industrial Technology Development Organization (NEDO).

### 4.1 Software and Statistical Analysis

We used the open-source TensorFlow library [8] to implement the models. We calculated the AUCs using the scikit-learn package [**?**] and plotted using matplotlib [9]. We computed the 95% CIs of the AUCs using the bootstrap method [10] with 1000 iterations.

## Declaration

The consent for publication is not applicable. Availability of Data and Material: The datasets generated and/or analysed during the current study are not publicly available due to specific institutional requirements governing privacy protection but are available from the corresponding author on reasonable request.

